# Estimating cancer risk in carriers of Lynch syndrome variants in UK Biobank

**DOI:** 10.1101/2023.11.10.23298308

**Authors:** Eilidh Fummey, Pau Navarro, John-Paul Plazzer, Ian M Frayling, Sara Knott, Albert Tenesa

## Abstract

**Background:** Lynch syndrome (LS) is an inherited cancer predisposition syndrome caused by genetic variants affecting DNA mismatch repair (MMR) genes *MLH1*, *MSH2*, *MSH6*, and *PMS2*. Cancer risk in LS is estimated from cohorts of individuals ascertained by family history of cancer, which is known to upwardly bias estimates.

**Methods:** The InSiGHT Database classifies MMR gene variants by pathogenicity through expert panel review of published evidence. 830 carriers of pathogenic or likely pathogenic (*path_MMR*) MMR gene variants from InSiGHT were identified in 454,756 UK Biobank participants using whole exome sequence. Nelson-Aalen survival analysis was used to estimate cumulative incidence of colorectal, endometrial, and breast cancer.

**Results:** Cumulative incidence of colorectal and endometrial cancer by age 70 was elevated in *path_MMR* carriers compared to non-carriers (colorectal: 11.8% (95% CI: 9.5 - 14.6) vs. 1.7% (1.6 - 1.7), endometrial: 13.4% (10.2 - 17.6) vs. 1.0% (0.9 - 1.0)), but the magnitude of this increase differed between genes. Cumulative breast cancer incidence by age 70 was not elevated in *path_MMR* carriers compared to non-carriers (8.9% (6.3 - 12.4) vs. 7.5% (7.4 - 7.6)). Cumulative cancer incidence estimates in UK Biobank were similar to estimates from the Prospective Lynch Syndrome Database for all genes and cancers, except there was no evidence for elevated endometrial cancer risk in carriers of pathogenic *PMS2* variants in UK Biobank.

**Conclusion:** These results can be used to inform the management of incidentally identified cases of LS. For example, they support the application of existing colorectal cancer surveillance strategies for LS in incidentally identified cases.

## Introduction

Lynch syndrome (LS), previously known as hereditary non-polyposis colorectal cancer (HNPCC), is an autosomal dominant cancer predisposition syndrome. LS is caused by variants affecting one of four DNA mismatch repair (MMR) genes: *MLH1*, *MSH2*, *MSH6*, or *PMS2*^1^. Individuals with LS generally have increased risk of developing early-onset colorectal cancer (CRC) and some extra-colonic cancers, such as endometrial cancer (EC). Understanding the cancer risk associated with likely pathogenic or pathogenic variants affecting MMR gene function (jointly referred to as *path_MMR*) is important for informing management plans for prevention and early detection of LS-associated cancers.

The majority of studies that estimate the cancer risk associated with *path_MMR* use cohorts of individuals who have a diagnosis of LS. These cohorts are subject to ascertainment bias because only individuals who have been diagnosed, or have a close family member who was diagnosed, with a LS-related cancer are likely to receive mutational screening. In other hereditary cancer predisposition syndromes, this has led to overestimation of cancer risk^2,3^.

If the cancer risk associated with *path_MMR* is overestimated, this poses a problem for counselling individuals with incidentally identified *path_MMR*. The American College of Medical Genetics and Genomics and Genomics England recommend reporting incidentally discovered *path_MMR* to patients even if they are asymptomatic and have no family history suggestive of LS^4,5^. Incidental identification of pathogenic variants is becoming more common because of large-scale biobanks that return genetic results to patients, like the National Institute of Health’s All of Us research program, and the increasing use of sequencing as a diagnostic tool in clinical genetics^6,7^.

Large population-based studies with sequencing data present an opportunity to estimate the penetrance of monogenic disease-causing variants without the bias introduced by selecting individuals based on disease status or family history^8^. However, these studies are potentially subject to healthy-volunteer bias and survivor bias, depending on the disease and study design, and thus may underestimate penetrance. Hence, such cohorts may provide a lower bound penetrance estimate, whereas classical studies of individuals diagnosed with rare genetic disease provide an upper bound.

The International Society for Gastrointestinal Hereditary Tumours (InSiGHT) maintains the Colon Cancer Gene Variant Database^9^. This database was created to bring together new and existing data on the clinical significance of germline variants for CRC risk. An expert panel reviews this evidence and classifies variants as pathogenic or benign following the International Agency for Research on Cancer’s five-tier classification system^10^.

Here, carriers of MMR gene variants listed in the InSiGHT Colon Cancer Gene Variant Database (herein referred to as the InSiGHT database) are identified in a large population-based study, UK Biobank (UKB), using whole exome sequence (WES) from 454,756 individuals. Carriers of *path_MMR* are used to generate cumulative incidence estimates for colorectal, endometrial, and breast cancer, and to test for the effect of sex and polygenic risk on CRC risk in LS. Additionally, cancer risk associated with variants of uncertain significance (VUS) is examined.

## Methods

### Phenotypic Data

UKB consists of ∼500,000 individuals living in the UK, recruited between the ages of 40 and 69^11^. At recruitment, individuals answered a health and lifestyle questionnaire, provided biological samples, and consented to their electronic health records, which include cancer and death registries, being linked to their UKB data.

The cancer registry data comes from two sources: NHS Digital for England and Wales, and the National Records of Scotland NHS Central Register for Scotland. For England and Wales, the data included recorded cancer cases from 1971 onwards and for Scotland, cases were recorded from 1957 onwards.

Only cancers that were diagnosed prior to the relevant cancer registry’s censor date were considered. The censor date is calculated by UKB, and is the last day of the month for which the number of records is greater than 90% of the mean number of records for the previous three months, unless the data is known to be incomplete, in which case the last day of the month before known incompleteness is used. The censoring dates for the data used here were 29/02/2020 for England and Wales and 31/01/2021 for Scotland. Death registry records were used to identify the appropriate end of observation for each participant.

The following ICD codes were used to identify relevant cancer diagnoses: ICD-9 153 – 154 and ICD-10 C18 – 20 for CRC; ICD-9 182 and ICD-10 C54 for EC; and ICD-9 174 and ICD-10 C50 for breast cancer (BC).

Family history of CRC was determined from recruitment questionnaire data. Participants were asked ’Has/did your [father/mother/brothers or sisters] ever suffered from any of the following illnesses?’, for which one of the responses was ’Bowel cancer’.

### Identification of InSiGHT Variant Carriers

Exome sequencing protocols in UKB have been described previously^12^. Genotypes supported by less than 10 reads or with a genotyping quality of less than 20 were set to missing. Sites were removed if more than 10% of genotypes were missing after these filters were applied, or if the site quality was less than 20. Variants located in exons 11 through 15 of *PMS2* were not considered due the homology of this region with pseudogene *PMS2CL*.

Chromosome, base pair position, reference, and alternative alleles for 1,932 InSiGHT classified variants were obtained from ClinVar and used to identify carriers from the UKB exome sequence. Variants are described relative to coding reference sequences on GRCh38.p14 (NC_000007.14): NM_000249.4(*MLH1*), NM_000251.2(*MSH2*), NM_000179.2(*MSH6*), and NM_000535.6(*PMS2*).

### Estimation of Cumulative Cancer Incidence

Cumulative cancer incidence was estimated using a modified form of Nelson-Aalen survival analysis as described in Dominguez-Valentin *et al.*^13^. Incidence rates were estimated for five-year intervals from age 25 to age 74, assuming no cancers occurred prior to the age of 25. The incidence rate within an interval is the number of events observed divided by the total number of years individuals were observed for within the interval, multiplied by five (the length of the interval). The cumulative incidences are obtained from incidence rates using the Nelson-Aalen estimator. Differences between survival curves were tested for using a Wilcoxon test where the proportional hazards assumption was not met and a log rank test otherwise.

### Polygenic Risk Score Calculation

Polygenic risk scores (PRS) for CRC were calculated for 694 *path_MMR* carriers who both self-identified as white British and had genetic principal components indicating European ancestry. The score was calculated as the sum of alleles at 76 variants as in Schmit *et al.*^14^, weighted by the effect estimates of the variants. The weights were calculated in samples independent of UKB. Genotypes used in PRS calculation were obtained from the genotyping array data available for UKB participants, described in Bycroft *et al.*^11^. Cox regression was used to test the effect of the PRS on CRC risk.

## Results

### 465 InSiGHT Variants Present in UKB

Amongst 454,756 UKB participants, 465 InSiGHT variants had at least one alternative allele carrier. Of these 465 variants, 160 were classified as benign or likely benign; 186 were variants of uncertain significance (VUS) and 119 were pathogenic or likely pathogenic (*path_MMR*).

The 119 different *path_MMR* variants were carried by 830 individuals (approximately 1 in 550 individuals). No individual carried more than one *path_MMR* and all individuals were heterozygous. Stratified by gene, 431 (51.9%) carried a pathogenic or likely pathogenic variant in *PMS2* (*path_PMS2*), 290 (34.9%) in *MSH6* (*path_MSH6*); 62 in *MSH1* (*path_MLH1*) (7.5%) and 47 in *MSH2* (*path_MSH2*) (5.7%).

Of the 119 *path_MMR* represented, 52 (43.7%) were observed in a single UKB participant (figure 1A). All but one *path_MMR* (PMS2:c.137G>T) was observed in 50 or fewer participants. An additional 7,849 UKB participants carried a VUS (approximately 1 in 60 individuals). The number of carriers per VUS ranged from one to 644 individuals (figure 1B).

**Figure 1:**
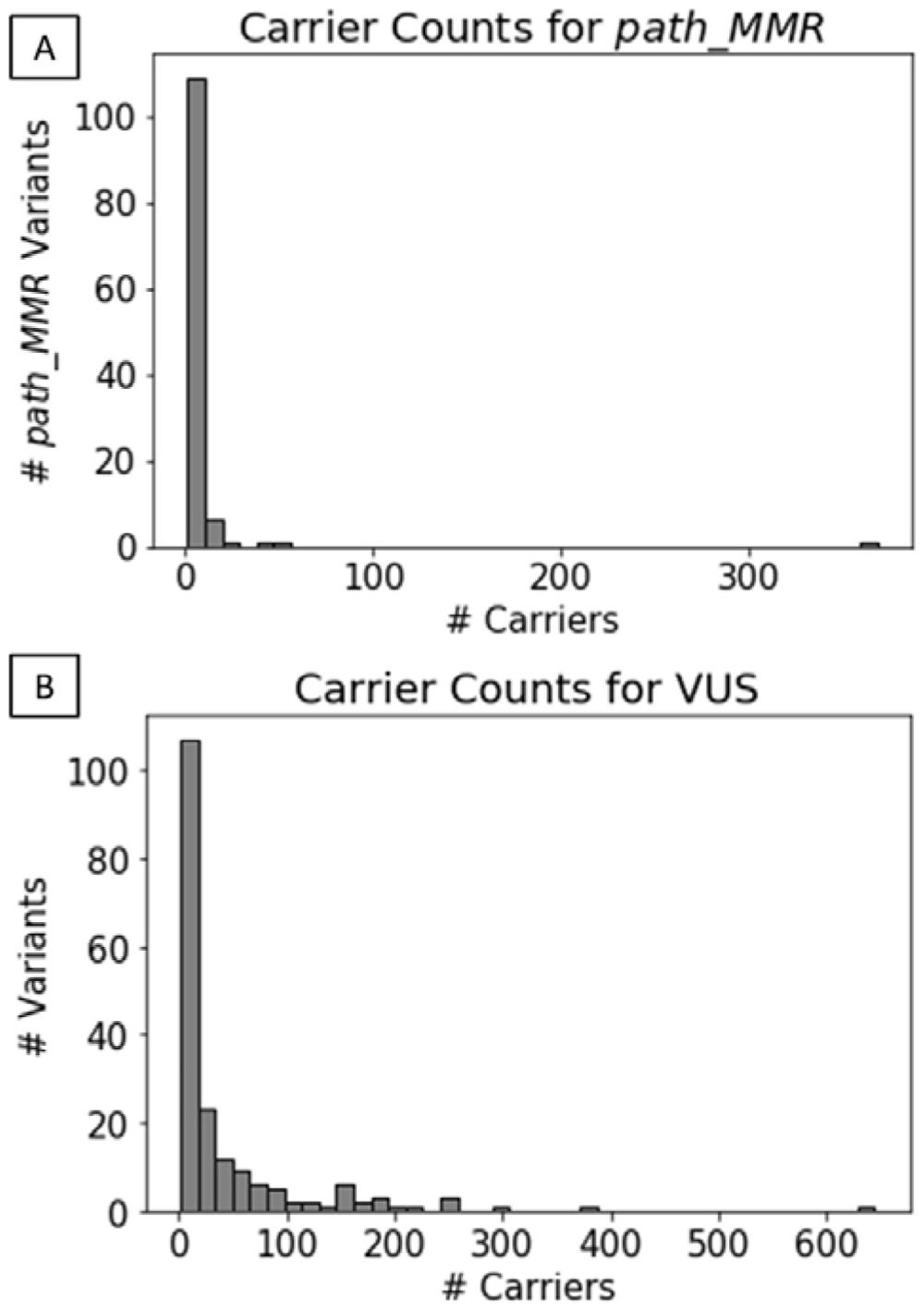
Histograms showing numbers of carriers in UK Biobank for MMR gene variants classified in the InSiGHT database as (A) pathogenic/likely pathogenic (*path_MMR*) and (B) of uncertain significance (VUS).

### Elevated Risk of Colorectal and Endometrial, but not Breast, Cancer in *path_MMR* **Carriers**

In total there were 7,854 participants with diagnoses of CRC, 84 (1.1%) of which occurred in *path_MMR* carriers, and 2,310 participants with diagnoses of EC, 54 (2.3%) of which occurred in *path_MMR* carriers. Nelson-Aalen curves for CRC and EC were fit for *path_MMR* carriers, stratified by gene, and for non-carriers. Leave-one-variant-out curves were generated for variants that made up more than 10% of carriers for a given gene. These curves were created using individuals who carried *path_MMR* for a given gene, excluding one particular variant, and were compared to the curve generated using all carriers of *path_MMR* for that gene to examine whether the shape of a curve for a gene was driven by a single variant. The leave-one-variant-out curves did not differ significantly from the all-variant curve: PMS2:c.137G>T (n = 368; Wilcoxon tests: colorectal: W = 0.05, p = 0.82; endometrial: W = 0.68, p = 0.41), MSH6:c.3226C>T (n = 50; log rank tests: colorectal: χ = 0.30, df = 1, p = 0.57; endometrial: χ = 0.47, df = 1, p = 0.49), and MSH6:c.3261dup (n = 46; Wilcoxon test, colorectal: W = 0.04, p = 0.84; log rank test, endometrial: χ^2^ = 0.25, df = 1, p = 0.62). This suggested that the results are not driven by a single variant.

The CRC Nelson-Aalen curve for *path_MMR* carriers was significantly different to the curve for non-carriers (log rank test: χ = 398.45, df = 1, p = 1.20 x 10), with cumulative incidence of CRC by age 70 elevated in *path_MMR* carriers compared to non-carriers (11.8% (95% CI: 9.5 - 14.6) vs. 1.7% (1.6 - 1.7)). Stratified by gene, *path_MMR* carriers had significantly different CRC curves from non-carriers at *p* < 0.01 (log rank tests: *path_MLH1*: χ^2^ = 882.63, df = 1, p = 5.86 x 10^-194^; *path_MSH2*: χ^2^ = 236.81, df = 1, p = 1.92 x 10^-53^; *path_MSH6*: χ^2^ = 182.88, df = 1, p = 1.14 x 10^-41^; *path_PMS2*: χ^2^ = 7.05, df = 1, p = 0.008).

In all five-year intervals, cumulative incidence of CRC for *path_MMR* carriers in all genes was equal to or greater than that of non-carriers (figure 2). Among *path_MMR* carriers, the cumulative incidence of CRC was highest for *path_MLH1* carriers and lowest for *path_PMS2* carriers in all intervals. The median age of CRC diagnosis was 51.8 for *path_MLH1* carriers; 52.3 for *path_MSH2*; 59.5 for *path_MSH6*; 67.2 for *path_PMS2* and 65.2 for individuals who do not carry *path_MMR*.

**Figure 2:**
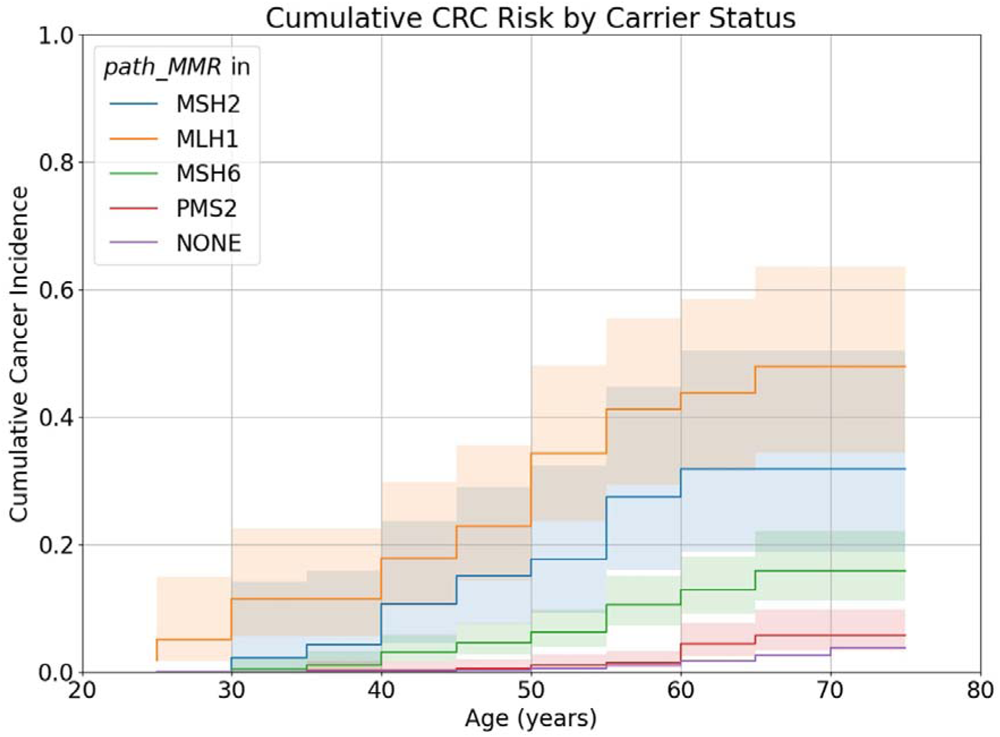
Nelson-Aalen curves of colorectal cancer for carriers of *path_MLH1*, *path_MSH2*, *path_MSH6*, *path_PMS2* and individuals who do not carry *path_MMR*. Shaded areas represent 95% CIs.

The EC Nelson-Aalen curve for *path_MMR* carriers was also significantly different to the curve for non-carriers (log rank test: χ = 660.85, df = 1, p = 9.76 x 10), with cumulative incidence of EC by age 70 elevated in *path_MMR* carriers compared to non-carriers (13.4% (95% CI: 10.2 - 17.6) vs. 1.0% (0.9 - 1.0)). When stratified by gene, only *path_MLH1*, *path_MSH2*, and *path_MSH6* carriers had significantly different EC survival curves from non-carriers at *p* < 0.01 (log rank tests: *path_MMR*: χ^2^ = 660.85, df = 1, p = 9.76 x 10^-146^; *path_MLH1*: χ^2^ = 470.93, df = 1, p = 2.01 x 10^-104^; *path_MSH2*: ^2^ = 95.6, df = 1, p = 1.14 x 10^-22^; *path_MSH6*: ^2^ = 935.13, df = 1, p = 2.27 x 10^-205^). In all five-year intervals, the cumulative EC incidence was greater for carriers of *path_MMR* variants in these genes than it was for non-carriers (figure 3). The median age of onset for *path_MLH1*, *path_MSH2*, and *path_MSH6* carriers was 56.4, 47.3, and 59.3, respectively, compared to 62.5 for non-carriers. However, *path_PMS2* carriers did not have a significantly different EC survival curve from non-carriers at p < 0.01 (log rank test; χ^2^ = 3.58, df = 1, p = 0.059).

**Figure 3:**
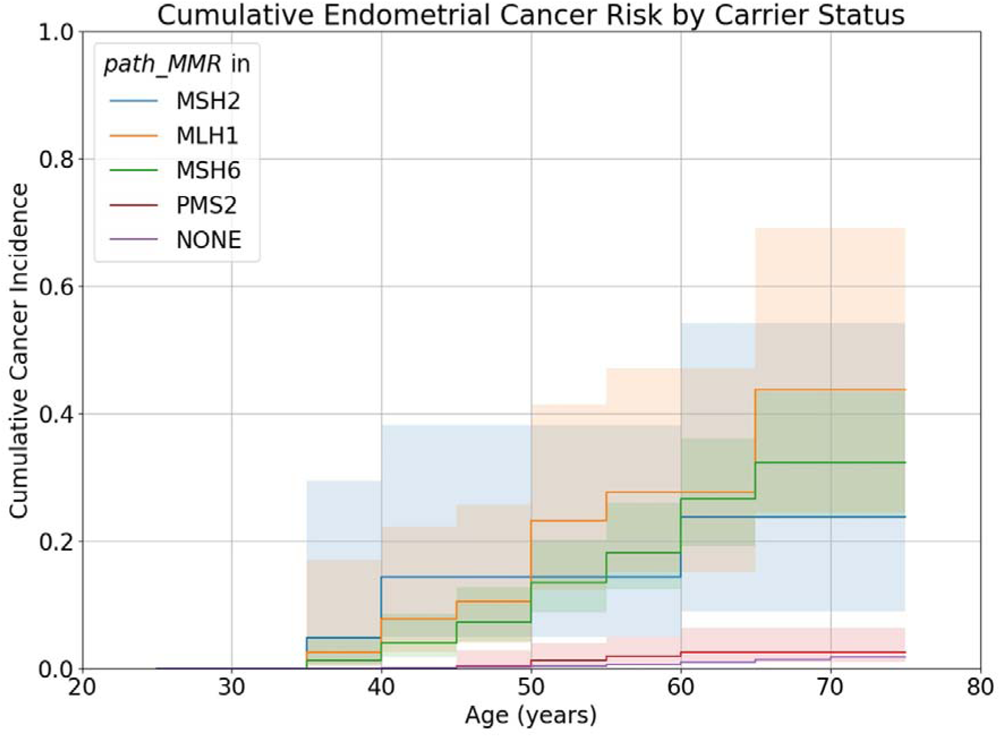
Nelson-Aalen curves of endometrial cancer for carriers of *path_MLH1*, *path_MSH2*, *path_MSH6*, *path_PMS2* and individuals who do not carry *path_MMR*. Shaded areas represent 95% CIs.

*path_MMR* carrier Nelson-Aalen curves for BC were not significantly different to the curve for non-carriers (Wilcoxon tests: *path_MMR*: W = 1.22, p = 0.27; *path_MLH1*: W = 0.21, p = 0.31; *path_MSH2*: W = 1.02, p = 0.31; *path_MSH6*: W = 0.12, p = 0.73; *path_PMS2*: W = 0.95, p = 0.33) (figure 4). Cumulative incidence of BC by age 70 in carriers of *path_MMR* was similar to that of non-carriers (8.9% (95% CI: 6.3 - 12.4) vs. 7.5% (7.4 - 7.6)).

**Figure 4:**
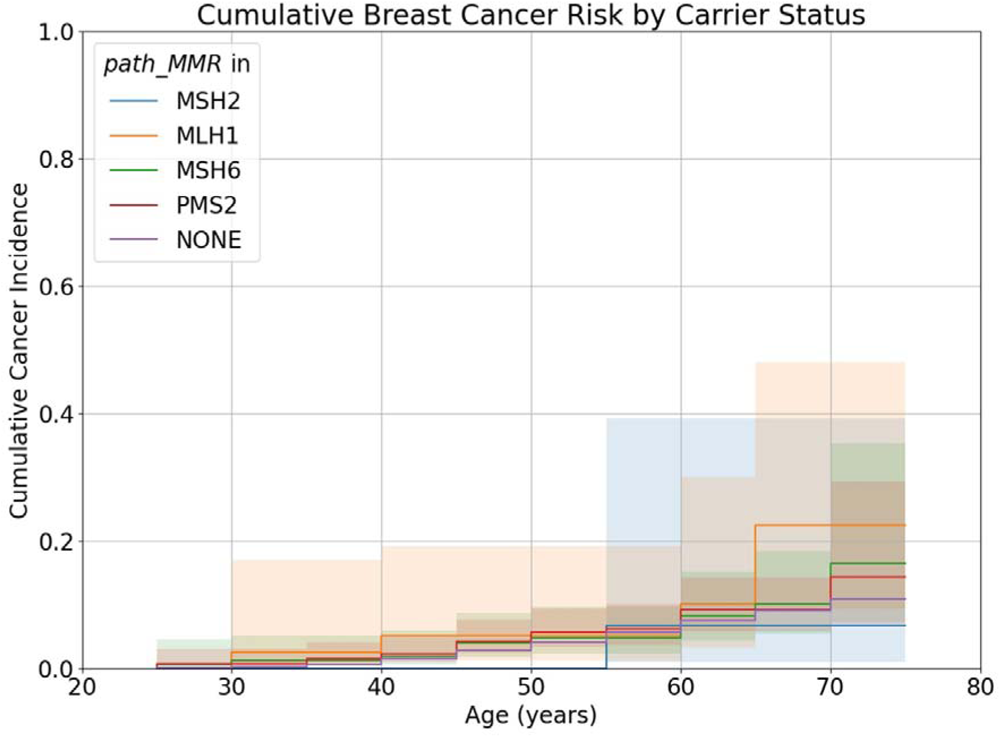
Nelson-Aalen curves of breast cancer for carriers of *path_MLH1*, *path_MSH2*, *path_MSH6*, *path_PMS2* and individuals who do not carry *path_MMR*. Shaded areas represent 95% CIs.

### Sex Differences in Colorectal Cancer Risk Observed for *path_MLH1* but Not Other ***path_MMR***

For each gene, CRC Nelson-Aalen curves of male and female *path_MMR* carriers were compared to establish whether sex influenced CRC risk (figure 5). The curve for male *path_MLH1* carriers was significantly different to the curve for female *path_MLH1* carriers at p < 0.05 (logrank test: χ^2^ = 4.29, df = 1, p = 0.038), although the 95% confidence intervals (CI) of both curves do intersect within all age intervals. Males had a higher cumulative incidence of CRC at age 70 (males: 53.5%, 95% CI: 34.7 – 74.7; females: 38.2%, 95% CI: 23.3 – 58.4) and an earlier median age of onset (males: 47.6; females: 55.1) (figure 5a). Differences between curves for males and females were not significant for any other gene (Wilcoxon tests: *path_MSH2*: W = 0.44, p = 0.50; *path_MSH6*: W = 0.15, p = 0.70; *path_PMS2*: W = 0.12, p = 0.73).

**Figure 5:**
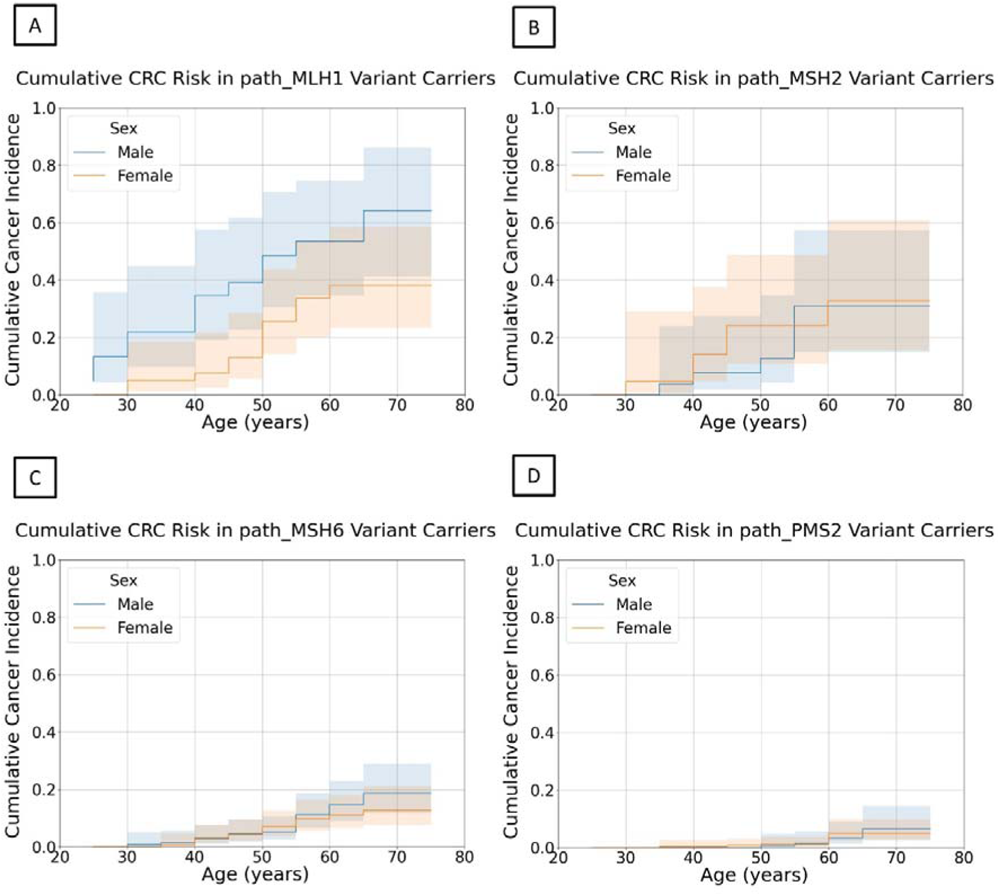
Nelson-Aalen curves of colorectal cancer for male and female carriers of (A) *path_MLH1*, (B) *path_MSH2*, (C) *path_MSH6*, and (D) *path_PMS2*. Shaded areas represent 95% CIs.

### *path_MMR* Carriers Report Family History of Colorectal Cancer More Frequently than Non-carriers

Higher proportions of *path_MMR* carriers reported a close family history (parent or sibling) of CRC than non-carriers (24.1% vs 10.4%), but the proportion differs significantly between MMR genes (chi square test: χ^2^ = 77.97, df = 3, p = 8.38 x 10^-17^). Over half of *path_MLH1* and *path_MSH2* carriers report a close family history of CRC (*path_MLH1*: 56.5%, *path_MSH2*: 51.1%). Reports of close family history of CRC were less frequent in *path_MSH6* carriers (27.2%) and even lower for *path_PMS2* carriers (14.4%). However, the proportion of close family history of CRC reported by *path_PMS2* carriers was still significantly greater than that of non-carriers (chi square test: χ = 6.76, df = 1, p = 0.0093).

### No Evidence that Polygenic Risk Modifies Colorectal Cancer Risk in *path_MMR* **Carriers**

Cox regression was used to test for the effect of the PRS developed in Schmit et al.^14^ on CRC risk within *path_MMR* carriers. White British UKB participants in the top 1% of this PRS had a 4-fold increased risk of CRC compared to individuals in the bottom 1%. For *path_MSH2*, *path_MSH6*, and *path_PMS2*, PRS was the sole predictor included in the Cox regression. For *path_MLH1*, the model included both sex and the PRS, as sex was a significant predictor at p < 0.05 (Wald test: b = 1.03, z = 2.40, p = 0.02). There is no evidence that the PRS modifies CRC risk for any gene (Wald test: *path_MLH1*: n = 51, b = 6.41, z = 1.62, p = 0.11; *path_MSH2*: n = 35, b = 1.47, z = 0.28, p = 0.78; *path_MSH6*: n = 242, b = 2.00, z = 0.64, p = 0.52; *path_PMS2*: n = 366, b = 3.98, z = 0.95, p = 0.34).

### Most Variants of Uncertain Significance Likely Benign

Sixty VUS were carried by 30 or more individuals. The proportion of CRC cases observed for carriers was greater than for non-carriers for 33 of these 60 VUS. None of the CRC Nelson-Aalen survival curves for these variants differed significantly from the non-carrier curve a p-value threshold, Bonferroni corrected for the number of VUS, of p < 0.0008. One variant had a curve that was significant at a nominal significance threshold of p < 0.05: MSH2:c.2400A>G (log rank test: χ^2^ = 8.80, df = 1, p = 0.003) (figure 6A). The cumulative incidence of CRC by age 70 in carriers of MSH2:c.2400A>G was 7.6% (95% CI: 3.7 – 15.3). This was lower than the cumulative incidence of CRC by age 70 for *path_MSH2* (31.8% (18.8 - 50.3)) although it was greater than *path_PMS2* (4.3% (2.4 - 7.6)). However, there was no evidence of increased family history of CRC in carriers of MSH2:c.2400A>G: 12.1% of carriers report a family history of CRC, compared to 10.5% of non-carriers (chi square test; χ^2^ = 0.32, df = 1, p = 0.57).

**Figure 6:**
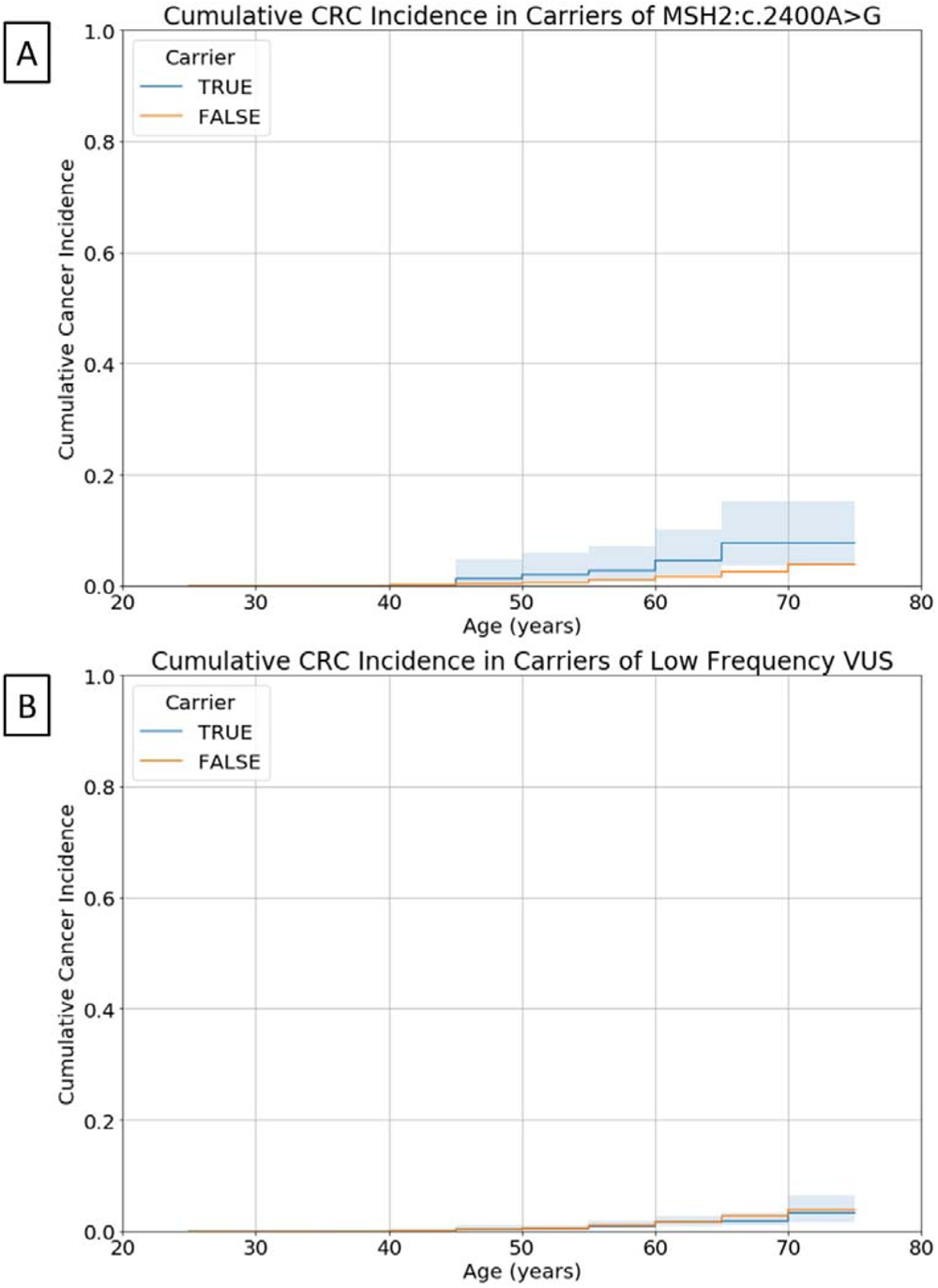
Nelson-Aalen curves of colorectal cancer for carriers (TRUE) of (A) the variant of uncertain significance, MSH2:c.2400A>G and (B) VUS with less than 30 carriers, compared to the curve for individuals who did not carry these variants (FALSE). Shaded areas represent 95% CIs.

Most *path_MMR* in UKB have less than 30 carriers (figure 1A). If pathogenic variants tend to be kept at low frequencies because of selection, it is possible that VUS that could not be tested individually due to an insufficient number of carriers will be the most likely to be pathogenic. There are 126 VUS with less than 30 carriers. A Nelson-Aalen curve for CRC was created for the 1,114 carriers of these low frequency VUS (figure 6B). This curve was not significantly different to that of participants that did not carry low frequency VUS (Wilcoxon test: W = 0.32, p = 0.57). Given this, it is likely that a majority of the low frequency VUS are also benign.

## Discussion

The analysis of carriers of InSiGHT classified *path_MMR* in UKB replicates known differences in the CRC and EC risk conferred by pathogenic variants in different MMR genes. EC risk is greater than CRC risk in *path_MSH6* carriers^13,15^. CRC risk is less and generally later onset in *path_MSH6* carriers than *path_MLH1* and *path_MSH2* carriers^13,15,16^. EC and CRC risk is notably less for *path_PMS2* carriers compared to carriers of *path_MMR* in other genes^13^.

In UKB, carriers of *path_MLH1* and *path_MSH2* are least frequently observed, many more *path_MSH6* carriers are identified, but the most carriers are observed for *path_PMS2*. This cannot be explained by greater numbers of InSiGHT classified *path_PMS2* variants compared to other genes, as there are more *path_MLH1* (535, 45.07%) and *path_MSH2* (454, 38.25%) variants than *path_MSH6* (167, 14.07%) or *path_PMS2* (31, 2.61%). Moreover, some regions of *PMS2* were not investigated due to pseudogene homology. The frequency of InSiGHT classified *path_MMR* in each gene in UKB is the inverse of the penetrance of these variants for CRC. This is expected if more penetrant alleles are subject to stronger purifying selection^17^.

In cohorts of individuals with LS ascertained from family history, the opposite trend is typically observed^15,18,19^. An example is Prospective Lynch Syndrome Database (PLSD), which collects data on carriers of *path_MMR* from the point of genetic diagnosis. At last publication, the database included 8,500 carriers: *path_MLH1* (36.8%) and *path_MSH2* (37.3%) carriers are most common, followed by *path_MSH6* (19.4%), and the fewest carriers are observed for *path_PMS2* (6.5%)^13^.

There are a number of potential explanations for this disparity between UKB and LS cohorts. Firstly, there are differences in how *path_MMR* were identified. The short read sequencing technology used to sequence UKB participants cannot accurately genotype structural variants, such as whole exon deletions or duplications, and thus these were not considered, although they are frequently identified in LS^20^. Additionally, only variants classified by InSiGHT as pathogenic or likely pathogenic were considered as *path_MMR* here to ensure sufficient weight of evidence for pathogenicity.

Cohort design may influence these differences in *path_MMR* carrier frequency. Given the early age of onset of cancers in *path_MLH1* and *path_MSH2* carriers, it is probable that they might be under-represented in UKB because they did not survive until recruitment age (40 - 69) or because individuals with health problems were less likely to respond to an invitation to participate^21,22^. *path_MSH6* and *path_PMS2* carriers may be under-represented in LS cohorts because lower penetrance obscures the autosomal dominant inheritance pattern used to identify affected families.

Despite differences in ascertainment between UKB and LS cohorts, CRC cumulative incidence estimates from UKB are similar to those in the literature^13,16,23^. Using PLSD as an example, the cumulative CRC incidence by age 70 for male carriers of *path_MLH1* is 53.5% (95% CI: 34.7 - 74.7) in UKB and 52.8% (46.1 – 59.8) in PLSD; for *path_MSH2*, 31.0% (14.9 – 57.32) vs 51.0% (42.8 – 59.7); for *path_MSH6*, 14.6% (9.1 – 23.0) vs 13.5% (7.1 – 24.8) and for *path_PMS2* 3.4% (1.3 – 9.0) vs 10.5% (2.7 – 36.0) (Figure S1). As in UKB, female carriers of *path_MLH1* in PLSD have a lower cumulative CRC incidence by age 70 than males: 42.1% (36.2 – 48.6) in UKB vs. 38.2% (23.25 – 58.39) in PLSD. Values are also similar for female carriers of *path_MMR* in the other three genes: for *path_MSH2,* 32.8% (15.5 – 60.7) in UKB vs. 39.8% (33.5 – 46.7) in PLSD; for *path_MSH6* 11.0% (6.6 – 18.1) vs 17.3% (11.2 – 26.3); and for *path_PMS2* 5.0% (2.5 – 9.9) vs 8.5% (2.1 – 31.5) (Figure S2).

However, it is unclear if *path_MMR* carriers in UKB have been diagnosed with LS, and are therefore receiving the same CRC surveillance offered to individuals in PLSD.

The cumulative EC incidence by age 70 for *path_MLH1* is 43.7% (95% CI: 24.5 – 69.1) in UKB vs 35.8% (29.9 – 42.5) in PLSD; for *path_MSH2* 23.7% (8.95 – 54.1) vs 42.1% (35.0 – 50.1); for *path_MSH6* 32.4% (23.5 – 43.5) vs 41.4% (32.3 – 52.0); and for *path_PMS2* 2.6% (1.07 – 6.32) vs 12.7% (5.5 – 27.9) (Figure S3). The only case where the 95% CI for UKB does not overlap the PLSD estimate and vice versa is for *path_PMS2*. In PLSD, carriers of *path_PMS2* have elevated EC risk but this trend is not observed in UKB.

Importantly, the similarity between UKB and PLSD does not suggest the existence of large subgroups of *path_MMR* carriers with low cancer risk that have been excluded from previous LS cancer risk estimates due to ascertainment bias. This is not the case in all monogenic diseases. One paper reported that the penetrance of diabetes in pathogenic *HNF1A*/*HNF4A* variant carriers was much lower in carriers identified through unselected cohorts (32%) versus cohorts of individuals with a molecular diagnosis (98%)^24^. However, there was not a difference in diabetes penetrance between different ascertainment contexts for carriers of pathogenic *GCK* variants. The data presented here for LS suggest that variants in all four of the MMR genes discussed are more similar to *GCK* than *HNF1A*/*HNF4A*: the method of identification of variant carriers does not change their penetrance estimates to such an extent that it would warrant different clinical management of incidentally identified carriers in most cases. The only exception is that *path_PMS2* carriers are observed to have lower EC risk in UKB than previously reported.

Elevated incidence of BC has been reported in individuals carrying *path_MSH6* and *path_PMS2*^25–27^. However, this finding is not consistently observed across all studies^23,28,29^. Elevated BC incidence is more commonly observed in studies where *path_MMR* carriers are identified in cohorts of individuals referred for non-specific hereditary cancer screening. It has been argued that because individuals with *path_MMR* are typically identified through screening of CRC patients, families with elevated risk of BC may be excluded, leading to inconsistent findings^26^.

If this were the case, we would expect to see elevated BC incidence in *path_MSH6* and *path_PMS2* carriers in UKB. Rather, there is no evidence of increased breast cancer risk in *path_MMR* carriers of any kind in UKB. Previous links between *path_MMR* and BC may have been caused by higher rates of mammographic screening in individuals with a family history of LS-associated cancers, such as ovarian, leading to overdiagnosis^23^. Additionally, due to their population frequencies, *path_MSH6* and *path_PMS2* may often be incidentally identified in individuals undergoing multigene hereditary cancer screening. However, the population undergoing this screening has a higher proportion of individuals with a BC diagnosis than the general population, so elevated BC risk in *path_MMR* carriers identified in this way could be attributed to ascertainment bias^29^.

One potential reason why one *path_MMR* carrier may develop CRC whilst another does not is additional genetic risk conferred by common variants. However, a PRS for CRC was not associated with CRC risk in UKB *path_MMR* carriers. This is consistent with previously published studies investigating the effects of common genetic variation on CRC risk in LS^30–32^. Although the power of these studies is limited by small sample sizes, it appears likely that the impact of common genetic variation on CRC risk in LS, if any, is too small to be useful for stratifying individuals into risk categories.

Around one in every sixty UKB participants carries a variant classified as of uncertain significance in the InSiGHT database. Thus, VUS are commonly encountered when performing mutational testing of MMR genes, introducing uncertainty around diagnosis. For most VUS observed in UKB, there was little evidence of pathogenicity. That most VUS behave like benign variants in UKB is consistent with the observation that VUS are more likely to be reclassified as benign or likely benign than upgraded to pathogenic or likely pathogenic^33–35^. Cumulative incidences of CRC for VUS are provided in Table S1. This evidence can be incorporated into future discussions about reclassification of VUS in MMR genes.

## Data Availability

Data can be obtained through application to UK Biobank.

https://www.ukbiobank.ac.uk/enable-your-research/apply-for-access.

