## supplementary_figures for "Estimating cancer risk in carriers of Lynch syndrome variants in UK Biobank"

Figure S1: Comparison between Nelson Aalen curves for colorectal cancer in UK Biobank (UKB, orange) and Prospective Lynch Syndrome Database (PLSD, blue) for male carriers of (A) *path_MLH1*, (B) *path_MSH2*, (C) *path_MSH6*, and (D) *path_PMS2*. Shaded areas represent 95% confidence intervals.

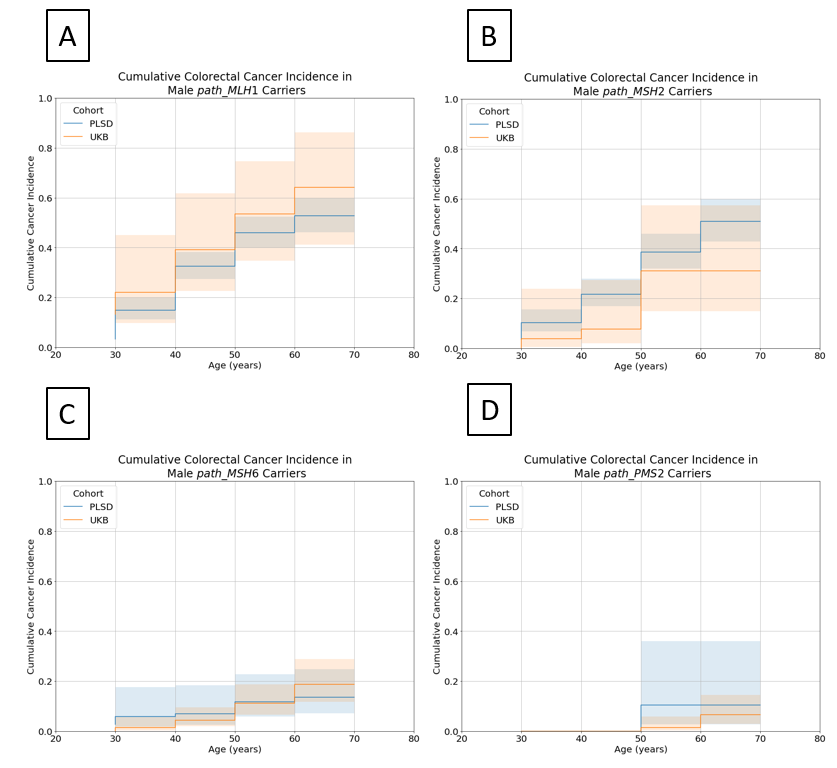

Figure S2: Comparison between Nelson Aalen curves for colorectal cancer in UK Biobank (UKB, orange) and Prospective Lynch Syndrome Database (PLSD, blue) for female carriers of (A) *path_MLH1*, (B) *path_MSH2*, (C) *path_MSH6*, and (D) *path_PMS2*. Shaded areas represent 95% confidence intervals.

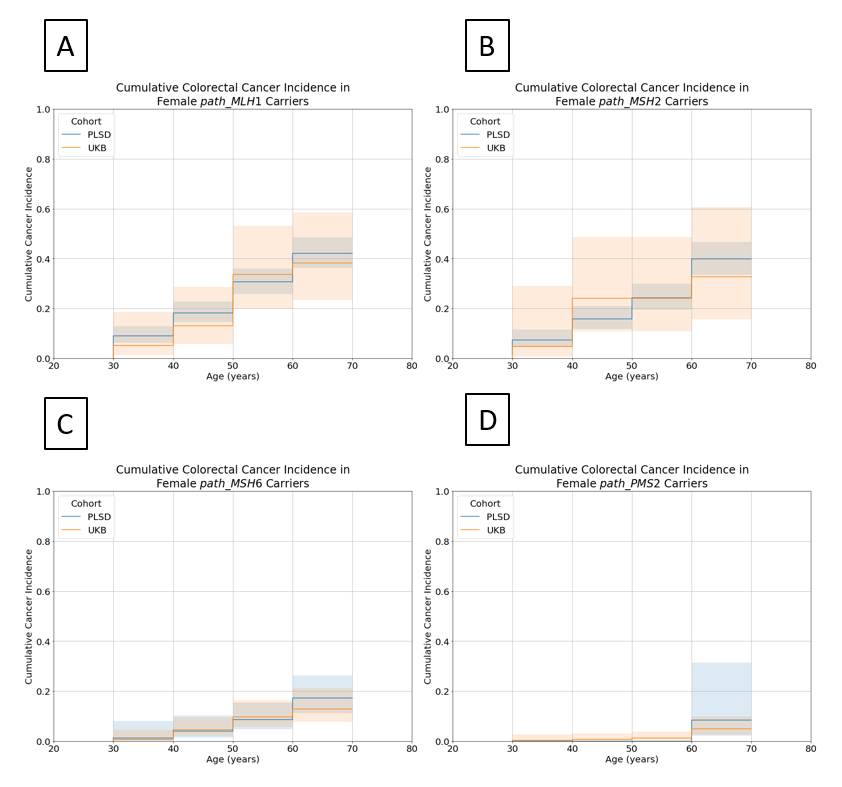

Figure S3: Comparison between Nelson Aalen curves for endometrial cancer in UK Biobank (UKB, orange) and Prospective Lynch Syndrome Database (PLSD, blue) for carriers of (A) *path_MLH1*, (B) *path_MSH2*, (C) *path_MSH6*, and (D) *path_PMS2*. Shaded areas represent 95% confidence intervals.

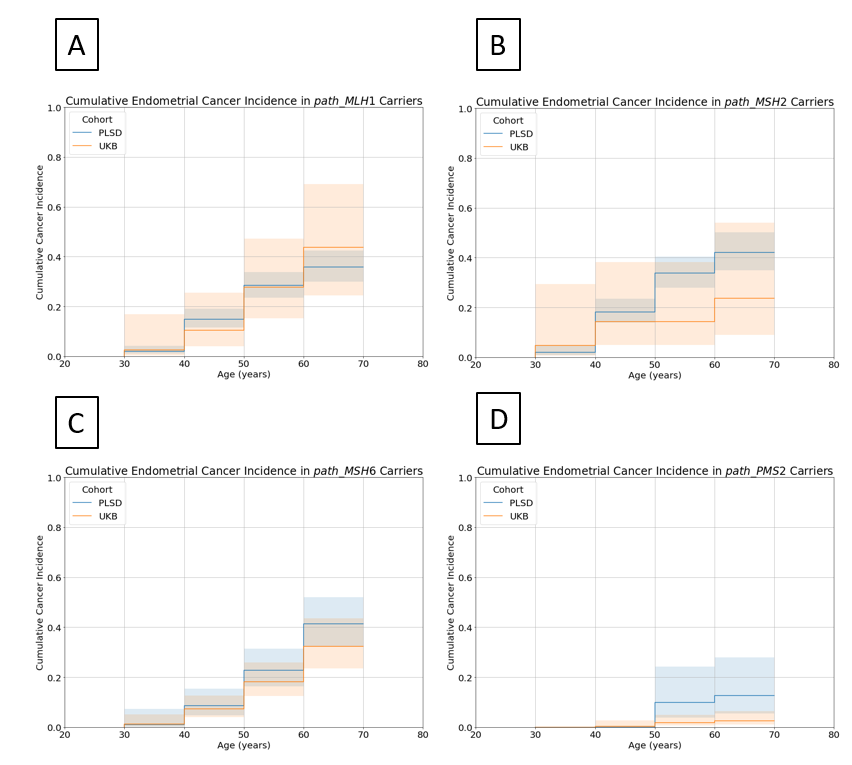
